# Psychosocial and socioeconomic vulnerability among caregivers of children with retinoblastoma: a cross-sectional latent profile study

**DOI:** 10.64898/2026.07.15.26358190

**Authors:** Peipei Zhang, Xiaohua Ge

## Abstract

**Background:** Caregivers of children with retinoblastoma (RB) face substantial psychological and socioeconomic challenges. However, the factors independently associated with caregiver burden and the distribution of risk across caregiver subgroups remain incompletely characterized. We examined psychosocial and socioeconomic correlates of caregiver burden, identified distinct vulnerability profiles, and evaluated factors associated with high-risk profile membership.

**Methods:** This cross-sectional study enrolled 413 primary caregivers of children with RB at a tertiary ophthalmic oncology center. Participants completed validated measures of caregiver burden (ZBI-22), anxiety (GAD-7), perceived social support (PSSS), family functioning (FAD-GF), and mental and physical quality of life (SF-12 MCS and PCS). Multivariable linear regression identified factors independently associated with caregiver burden and mental quality of life. Mediation analysis evaluated the indirect association between social support and burden through family functioning, and moderation analysis assessed whether household income modified the association between family dysfunction and burden. Latent profile analysis (LPA) identified caregiver risk profiles, and multinomial logistic regression examined factors associated with profile membership.

**Results:** Anxiety showed the strongest independent association with greater caregiver burden (standardized coefficient beta = 0.641, 95% CI [1.46, 1.84], P < 0.001) and poorer mental quality of life (beta = −0.483, 95% CI [−0.12, −0.08], P < 0.001). Family debt was independently associated with greater burden (beta = 0.195, P = 0.040). Family functioning accounted for 32.19% of the total association between social support and burden. Household income modified the association between family dysfunction and burden (interaction B = −0.85, P < 0.001), with a steeper gradient in lower-income households. LPA identified three profiles: severe burden-high vulnerability (n = 82, 19.85%), moderate burden (n = 193, 46.73%), and mild burden-high resilience (n = 138, 33.41%). Low-to-moderate household income was associated with higher odds of severe-profile membership (OR = 31.50, 95% CI [6.56, 151.24], P < 0.001).

**Conclusions:** Caregiver burden in pediatric RB was associated more strongly with psychosocial and socioeconomic factors than with the clinical indicators examined. Family functioning partly accounted for the association between social support and burden, while household income modified the association between family dysfunction and burden. These findings support prospective evaluation of family-centered and financial-support interventions and suggest that profile-based screening may help identify caregivers requiring more intensive support.

**Key Messages:** *What is already known on this topic:* Caregivers of children with retinoblastoma may experience psychological distress and financial strain related to visual loss, enucleation, prolonged treatment and repeated surveillance. However, caregiver heterogeneity and the joint contribution of psychosocial and socioeconomic factors remain insufficiently characterised.

*What this study adds:* In 413 caregivers, anxiety showed the strongest association with caregiver burden, while lower household income amplified the association between family dysfunction and burden. Latent profile analysis identified a high-burden, high-vulnerability subgroup comprising approximately one-fifth of caregivers, in whom financial hardship was concentrated.

*How this study might affect research, practice or policy:* Multidisciplinary retinoblastoma services may benefit from prospective evaluation of caregiver distress, family functioning and financial-risk screening, particularly during treatment-intensive periods.

## Introduction

Retinoblastoma (RB) is the most common primary intraocular malignancy of childhood, with approximately 8,000 new cases diagnosed worldwide each year [1]. Multidisciplinary treatment has substantially improved survival in high-income settings [2]. However, treatment may involve enucleation, systemic or intra-arterial chemotherapy, and repeated surveillance examinations under anesthesia, placing sustained psychological and financial demands on family caregivers [3]. Caregivers must contend with the threat of vision loss or death, repeated hospital visits, treatment-related financial strain, and decisions about irreversible interventions such as eye removal [4].

Previous studies have reported anxiety, depression, and impaired quality of life among caregivers of children with RB [5], together with substantial out-of-pocket costs [3]. Two limitations remain. First, most studies have examined individual risk factors separately rather than evaluating psychological distress, social support, family functioning, and economic hardship within one analytical framework. Second, population-level estimates may conceal clinically relevant heterogeneity because caregivers can experience markedly different combinations of burden and vulnerability.

We therefore assessed anxiety, perceived social support, family functioning, household income, and family debt within a single analytical framework. We also used latent profile analysis (LPA) to complement variable-centered analyses with a person-centered description of caregiver heterogeneity. This approach was intended to identify vulnerable caregiver subgroups and potentially modifiable factors associated with their risk.

Cross-disease comparisons suggest that the mean caregiver burden in the present RB cohort (ZBI-22 = 26.50) was lower than values reported for cerebral palsy (36.31), pediatric leukemia (33.15), and neurofibromatosis type 1 (34.31) [6–8]. However, the severe-burden RB profile had a mean ZBI-22 score of 46.41, approaching the value reported among uninsured families of children receiving active cancer treatment (48.66) [9]. These comparisons indicate that cohort averages may obscure a substantially distressed subgroup and support person-centered risk stratification.

Accordingly, this cross-sectional study aimed to (1) identify factors independently associated with caregiver burden and mental quality of life; (2) evaluate statistical pathways linking social support, family functioning, household income, and caregiver burden; and (3) classify caregivers into distinct profiles and quantify factors associated with high-risk profile membership.

## Methods

### Study Design and Participants

This cross-sectional study was conducted at a tertiary ophthalmic oncology center in China and was reported in accordance with the Strengthening the Reporting of Observational Studies in Epidemiology (STROBE) guidelines [10]. The institutional review board approved the study, and all participants provided written informed consent. We consecutively enrolled 413 primary family caregivers of children diagnosed with RB during clinic visits. Eligibility criteria were (1) being the primary caregiver of a child with RB, (2) caring for a child undergoing active treatment or follow-up, and (3) being able to provide informed consent. Sample size planning used G*Power 3.1 [11], assuming a medium effect size (f2 = 0.15), 80% power, and a two-sided alpha of 0.05 for multiple regression.

### Measures

Caregivers completed validated questionnaires. Caregiver burden was assessed with the 22-item Zarit Burden Interview (ZBI-22; range, 0-88), with higher scores indicating greater burden [12]. The Chinese ZBI-22 has demonstrated good internal consistency (Cronbach alpha = 0.88) [13]. Anxiety was measured with the 7-item Generalized Anxiety Disorder scale (GAD-7), which has shown high reliability in primary care populations (Cronbach alpha = 0.92) [14]. Perceived social support was measured with the Perceived Social Support Scale (PSSS), a Chinese adaptation [15] of the Multidimensional Scale of Perceived Social Support [16]. Family functioning was assessed with the 12-item General Functioning subscale of the Family Assessment Device (FAD-GF), for which higher scores indicate greater family dysfunction [17]. Mental and physical quality of life were measured using the mental and physical component summaries of the 12-item Short Form Health Survey (SF-12 MCS and PCS) [18]. Economic indicators included annual post-diagnosis household income, categorized into six ordinal levels, and treatment-related family debt (yes/no). Clinical variables included disease laterality and enucleation status.

### Statistical Analysis

All quantitative analyses were performed using IBM SPSS Statistics version 27.0 [19] and R version 4.2.3 [20]. All tests were two-sided, with alpha = 0.05, and effect estimates were reported with 95% confidence intervals where available.

#### Descriptive and bivariate analyses

Continuous variables are presented as means and standard deviations, and categorical variables as frequencies and percentages. Independent-samples t tests were used for two-group comparisons, and one-way analysis of variance was used for comparisons involving three or more groups. Spearman correlations were used to evaluate bivariate associations among continuous variables.

#### Multivariable linear regression

Factors independently associated with ZBI-22 and SF-MCS-12 scores were identified using bidirectional stepwise selection based on the Akaike information criterion.

Standardized coefficients (beta) were reported to facilitate comparison of effect magnitudes. Collinearity was assessed using variance inflation factors, with values below 5 considered acceptable.

#### Mediation and moderation analyses

We used the PROCESS macro [21] with 5,000 bootstrap resamples. Model 4 evaluated whether FAD-GF statistically mediated the association between PSSS and ZBI-22.

Model 1 evaluated whether household income moderated the association between FAD-GF and ZBI-22. Conditional effects were estimated at 1 SD below the mean, the mean, and 1 SD above the mean of the moderator.

#### Latent profile analysis

LPA was performed with the tidyLPA package in R [22] using standardized GAD-7, PSSS, and ZBI-22 scores as indicators. Models with one to five profiles were compared using the Akaike information criterion, Bayesian information criterion, sample-size-adjusted Bayesian information criterion, entropy, Lo-Mendell-Rubin likelihood ratio test, bootstrap likelihood ratio test, and interpretability [23]. Distal outcomes (FAD-GF, SF-MCS-12, and SF-PCS-12) were compared across profiles using analysis of variance.

Multinomial logistic regression. Factors associated with LPA profile membership were examined using multinomial logistic regression, with the mild-burden profile as the reference category. Adjusted odds ratios (ORs) and 95% confidence intervals (CIs) are reported.

## Results

### The Vulnerability Profile of Retinoblastoma Caregivers

The analysis included 413 primary family caregivers of children with RB (Table 1). Most caregivers were female (293/413, 70.9%), and 295 households (71.4%) had accumulated treatment-related debt.

**Table 1.** Baseline characteristics and univariate analysis of caregiver burden (N = 413). Data are presented as n (%) for categorical variables and mean +/−SD for ZBI-22 scores. Between-group differences were evaluated using independent-samples t tests for two-group comparisons and one-way analysis of variance for comparisons involving three or more groups. All tests were two-sided. ZBI-22, 22-item Zarit Burden Interview.

| Characteristics | N (%) | ZBI Score (Mean ± SD) | Statistic (t/F) | P value |
| --- | --- | --- | --- | --- |
| **Caregiver Gender** |  |  | t = 1.13 | 0.258 |
| Female | 293 (70.9%) | 27.03 ± 15.07 | | |
| Male | 120 (29.1%) | 25.20 ± 14.86 | | |
| **Caregiver Education Level** |  |  | F = 3.69 | 0.006 |
| Associate degree | 95 (23.0%) | 28.29 ± 13.98 | | |
| Bachelor degree | 121 (29.3%) | 23.68 ± 14.16 | | |
| High school | 93 (22.5%) | 24.98 ± 14.26 | | |
| Master or above | 22 (5.3%) | 24.09 ± 12.50 | | |
| Middle school or below | 82 (19.9%) | 30.96 ± 17.66 | | |
| **Family Debt** |  |  | t = -7.18 | < 0.001 |
| No | 118 (28.6%) | 19.57 ± 10.91 | | |
| Yes | 295 (71.4%) | 29.27 ± 15.55 | | |
| **Child Gender** |  |  | t = 1.76 | 0.080 |
| Boy | 233 (56.4%) | 27.64 ± 15.20 | | |
| Girl | 180 (43.6%) | 25.03 ± 14.69 | | |
| **Onset Type** |  |  | t = 2.29 | 0.023 |
| Bilateral | 133 (32.2%) | 28.99 ± 15.50 | | |
| Unilateral | 280 (67.8%) | 25.32 ± 14.66 | | |
| **Eye Enucleation** |  |  | t = 0.89 | 0.376 |
| No | 39 (9.4%) | 28.67 ± 16.00 | | |
| Yes | 374 (90.6%) | 26.28 ± 14.92 | | |
| **Post-illness Income** |  |  | F = 10.69 | < 0.001 |
| 1 | 77 (18.6%) | 35.14 ± 16.25 | | |
| 2 | 121 (29.3%) | 26.73 ± 13.79 | | |
| 3 | 113 (27.4%) | 25.64 ± 14.57 | | |
| 4 | 63 (15.3%) | 23.54 ± 13.19 | | |
| 5 | 25 (6.1%) | 17.32 ± 10.42 | | |
| 6 | 14 (3.4%) | 13.71 ± 12.63 | | |

Caregiver burden varied more strongly across socioeconomic characteristics than across the clinical characteristics examined. The largest bivariate difference was between caregivers with and without family debt (mean ZBI-22, 29.27 +/−15.55 vs. 19.57 +/−10.91; t = −7.18, P < 0.001). Burden decreased across post-illness household-income levels, from 35.14 +/−16.25 in level 1 to 13.71 +/−12.63 in level 6 (F = 10.69, P < 0.001; Figure 1C). Burden also differed across daily-care-hour categories (F = 10.27, P < 0.001; Figure 1E). Bilateral disease was associated with greater burden than unilateral disease (28.99 +/−15.50 vs. 25.32 +/−14.66; t = 2.29, P = 0.023), whereas caregiver gender and enucleation status were not associated with burden (P = 0.258 and P = 0.376, respectively).

**Figure 1.**
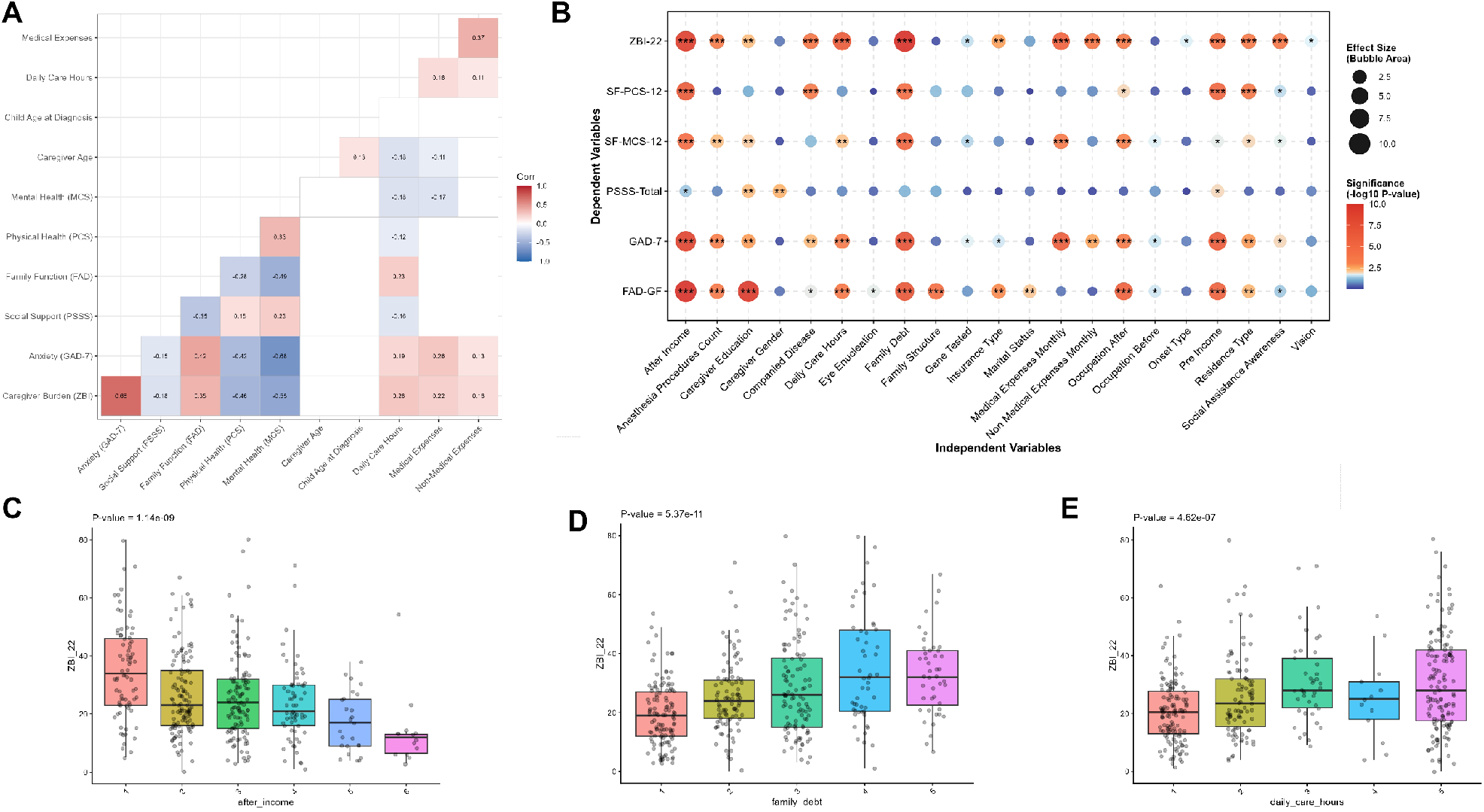
Landscape of caregiver burden and bivariate associations. (A) Spearman correlation heatmap showing associations among the principal continuous variables. Red denotes positive correlations and blue denotes negative correlations. The strongest correlation was between caregiver burden (ZBI-22) and anxiety (GAD-7) (r = 0.68, P < 0.001). (B) Bubble matrix showing statistical significance (−log10 P value) and effect size (bubble area) for associations between independent variables and key outcomes. Red indicates smaller P values, and bubble area represents effect magnitude. Asterisks denote statistical significance (*P < 0.05, **P < 0.01, ***P < 0.001). (C-E) Boxplots showing caregiver burden across post-diagnosis household-income levels (C; F = 10.69, P < 0.001), family-debt status (D; t = −7.18, P < 0.001), and daily-care-hour categories (E; F = 10.27, P < 0.001). Individual observations are overlaid. Group differences were evaluated using one-way analysis of variance (C,E) or an independent-samples t test (D). Abbreviations: FAD-GF, Family Assessment Device-General Functioning; GAD-7, Generalized Anxiety Disorder-7; PSSS, Perceived Social Support Scale; SF-MCS-12, Short Form-12 Mental Component Summary; SF-PCS-12, Short Form-12 Physical Component Summary; ZBI, Zarit Burden Interview.

Anxiety was strongly correlated with caregiver burden (r = 0.68, P < 0.001; Figure 1A), representing the largest bivariate association among the continuous variables examined (Figure 1B). These findings informed the subsequent multivariable analyses.

### Independent Drivers of Caregiver Burden and Quality of Life

Multivariable linear regression models were used to estimate the independent associations of psychosocial and socioeconomic factors with caregiver burden and mental quality of life (Table 2; Figure 2A,B).

**Table 2.** Multivariable linear regression models for caregiver burden and mental quality of life. Final models were selected using bidirectional stepwise selection based on the Akaike information criterion. B, unstandardized coefficient; SE, standard error; beta, standardized coefficient. Panel A (dependent variable: ZBI-22 caregiver burden): adjusted R2 = 0.531, F(8,404) = 53.70, P < 0.001. Panel B (dependent variable: SF-MCS-12 mental quality of life): adjusted R2 = 0.554, F(9,403) = 58.78, P < 0.001. Residence type was entered as a five-level categorical variable. CI, confidence interval.

| Variables | B | SE | $\beta$ | 95% CI | P value |
| --- | --- | --- | --- | --- | --- |
| **Panel: Panel A: Caregiver Burden (ZBI)** |  |  |  |  |  |
| Anxiety (GAD-7) | 1.65 | 0.10 | 0.641 | 1.46, 1.84 | < 0.001 |
| Social Support (PSSS) | -0.10 | 0.06 | -0.066 | -0.21, 0.01 | 0.071 |
| Daily Care Hours | 1.09 | 0.38 | 0.119 | 0.35, 1.83 | 0.004 |
| Monthly Non-medical Expenses | 0.74 | 0.53 | 0.052 | -0.29, 1.78 | 0.159 |
| Pre-illness Income | 1.54 | 0.94 | 0.116 | -0.30, 3.38 | 0.100 |
| Post-illness Income | -2.00 | 0.89 | -0.167 | -3.75, -0.26 | 0.025 |
| Caregiver Education Level | 1.03 | 0.54 | 0.082 | -0.02, 2.09 | 0.056 |
| Family Debt (Ref: No) | 2.97 | 1.44 | 0.195 | 0.14, 5.81 | 0.040 |
| **Panel: Panel B: Quality of Life (SF-MCS-12)** |  |  |  |  |  |
| Anxiety (GAD-7) | -0.10 | 0.01 | -0.483 | -0.12, -0.08 | < 0.001 |
| Social Support (PSSS) | 0.01 | 0.00 | 0.079 | 0.00, 0.02 | 0.033 |
| Family Function (FAD-GF) | -0.57 | 0.12 | -0.197 | -0.80, -0.34 | < 0.001 |
| Residence Type [2] | 0.68 | 0.30 | 0.546 | 0.09, 1.27 | 0.023 |
| Residence Type [3] | 0.47 | 0.29 | 0.373 | -0.11, 1.04 | 0.115 |
| Residence Type [4] | 0.69 | 0.29 | 0.555 | 0.12, 1.27 | 0.018 |
| Residence Type [5] | 0.73 | 0.29 | 0.582 | 0.15, 1.30 | 0.013 |
| Caregiver Burden (ZBI-22) | -0.01 | 0.00 | -0.180 | -0.02, -0.01 | < 0.001 |

**Figure 2.**
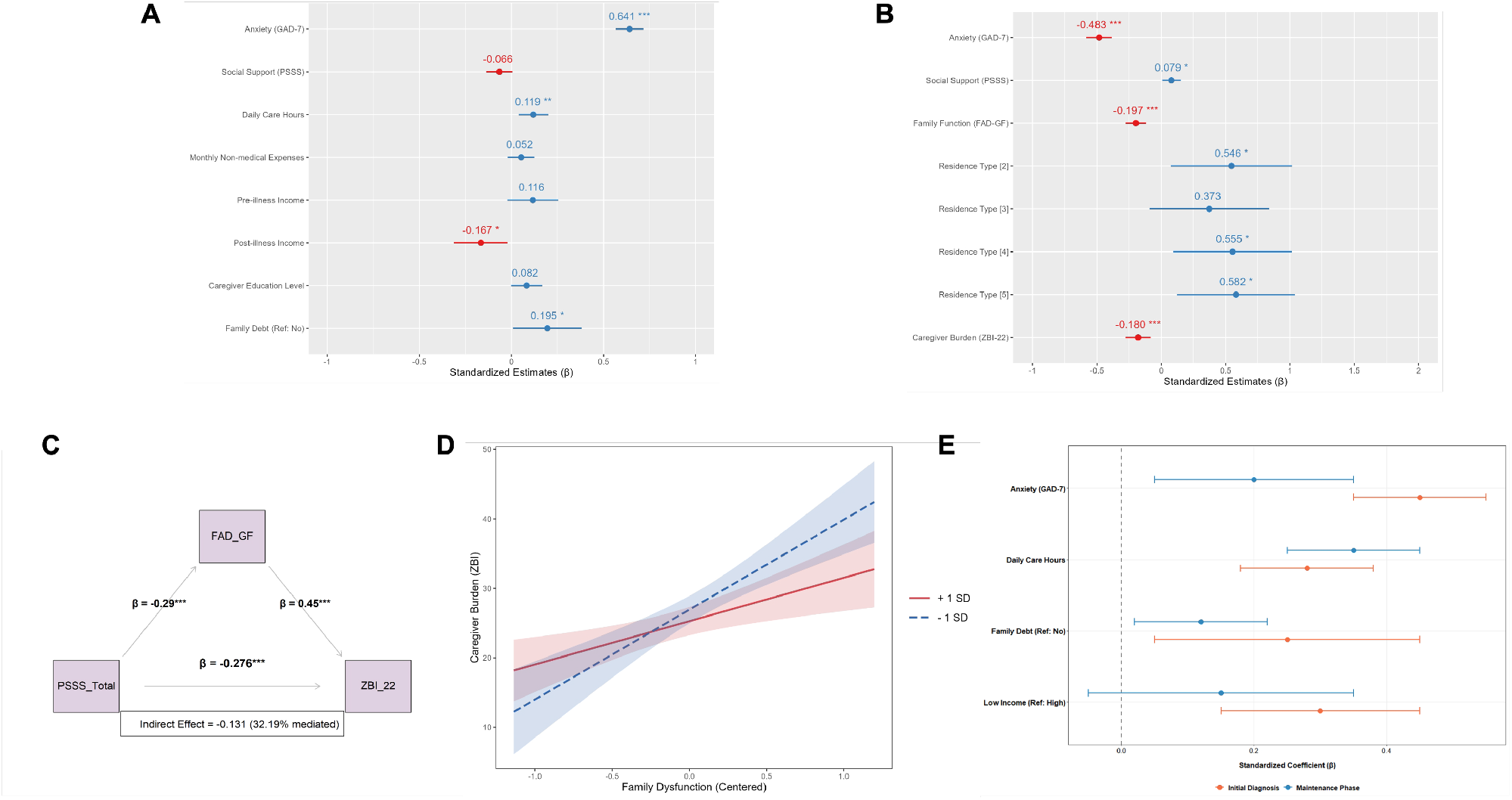
Mechanistic pathways of caregiver burden: independent predictors, mediation, and moderation. (A) Forest plot of standardized coefficients (beta) and 95% CIs from the multivariable model of caregiver burden (adjusted R2 = 0.531, F = 53.70, P < 0.001). Anxiety had the largest positive standardized coefficient (beta = 0.641, P < 0.001). Blue denotes positive associations and red denotes negative associations. (B) Forest plot of predictors in the model of mental quality of life (adjusted R2 = 0.554, F = 58.78, P < 0.001). Anxiety had the largest negative standardized coefficient (beta = −0.483, P < 0.001). (C) Path diagram showing partial statistical mediation by family functioning in the association between perceived social support and caregiver burden. The indirect association accounted for 32.19% of the total association (beta = −0.131, 95% bootstrap CI [−0.184, −0.089], P < 0.001). (D) Conditional-effects plot showing moderation by household income of the association between family dysfunction and caregiver burden (interaction B = −0.85, P < 0.001). Slopes are shown at lower income (−1 SD; 3.30, P < 0.001), mean income (2.45, P < 0.001), and higher income (+1 SD; 1.60, P = 0.002). Shaded bands denote 95% CIs. (E) Forest plot comparing standardized coefficients for anxiety, daily care hours, family debt, and lower income between the initial-diagnosis and maintenance phases. Error bars denote 95% CIs. Statistical significance is indicated by *P < 0*.*05, P < 0*.*01*, **P < 0.001. Abbreviations: CI, confidence interval; SD, standard deviation**.

In the caregiver-burden model (Table 2, Panel A), anxiety had the largest standardized coefficient (beta = 0.641, 95% CI [1.46, 1.84], P < 0.001). Family debt (beta = 0.195, 95% CI [0.14, 5.81], P = 0.040) and daily care hours (beta = 0.119, 95% CI [0.35, 1.83], P = 0.004) were independently associated with greater burden. Higher post-illness household income was associated with lower burden (beta = −0.167, 95% CI [−3.75, −0.26], P = 0.025). The model explained 53.1% of the variance in caregiver burden (adjusted R2 = 0.531, F[8,404] = 53.70, P < 0.001).

In the mental-quality-of-life model (Table 2, Panel B), anxiety had the largest negative standardized coefficient (beta = −0.483, 95% CI [−0.12, −0.08], P < 0.001). Family dysfunction (beta = −0.197, 95% CI [−0.80, −0.34], P < 0.001) and caregiver burden (beta = −0.180, 95% CI [−0.02, −0.01], P < 0.001) were also negatively associated with SF-MCS-12 scores. Perceived social support showed a modest positive association (beta = 0.079, 95% CI [0.00, 0.02], P = 0.033). The model explained 55.4% of the variance (adjusted R2 = 0.554, F[9,403] = 58.78, P < 0.001). Among the variables reported in Supplementary Table S1, all variance inflation factors were below 5.

Across both models, anxiety had the largest standardized coefficient. Disease laterality and enucleation status were not retained in either final model. The bivariate association between social support and burden was attenuated after adjustment, suggesting overlap with psychological, family, and economic factors. The mediation analysis therefore evaluated family functioning as one potential statistical pathway.

### Mechanistic Pathways: Mediation by Family Functioning and Moderation by Income

Mediation analysis evaluated whether family functioning statistically accounted for part of the association between perceived social support and caregiver burden (Figure 2C; Supplementary Table S2). The total association was significant (beta = −0.407, 95% CI [−0.518, −0.301], P < 0.001). The indirect association through family functioning was also significant (beta = −0.131, 95% bootstrap CI [−0.184, −0.089], P < 0.001) and represented 32.19% of the total association. The direct association remained significant after inclusion of the mediator (beta = −0.276, 95% CI [−0.390, −0.158], P < 0.001), consistent with partial statistical mediation.

Moderation analysis assessed whether household income modified the association between family dysfunction and caregiver burden (Figure 2D; Supplementary Table S3). The interaction was significant (B = −0.85, 95% CI [−1.09, −0.61], P < 0.001). The estimated slope was greatest at lower income (3.30, P < 0.001), decreased at the mean income level (2.45, P < 0.001), and was smallest at higher income (1.60, P = 0.002). Thus, the association between family dysfunction and burden was approximately twice as steep at lower income as at higher income.

Phase-stratified analyses indicated stronger associations of family debt and lower income with caregiver burden during the initial-diagnosis phase than during the maintenance phase (Figure 2E).

### Clinical Stratification: Latent Profiles and Risk Prediction

LPA identified three caregiver profiles based on anxiety, perceived social support, and caregiver burden (Figure 3A). Profile A (severe burden-high vulnerability; n = 82, 19.85%) had high anxiety (GAD-7, 16.78), high burden (ZBI-22, 46.41), low social support (PSSS, 50.45), greater family dysfunction (FAD-GF, 2.42), and the lowest mean income level (2.54). Profile C (mild burden-high resilience; n = 138, 33.41%) showed low anxiety (1.61), low burden (14.94), higher social support (54.52), less family dysfunction (1.94), and the highest mean income level (3.38). Profile B (moderate burden; n = 193, 46.73%) had intermediate values (ZBI-22, 26.31; GAD-7, 8.17; PSSS, 50.47; FAD-GF, 2.16; income, 2.95).

**Figure 3.**
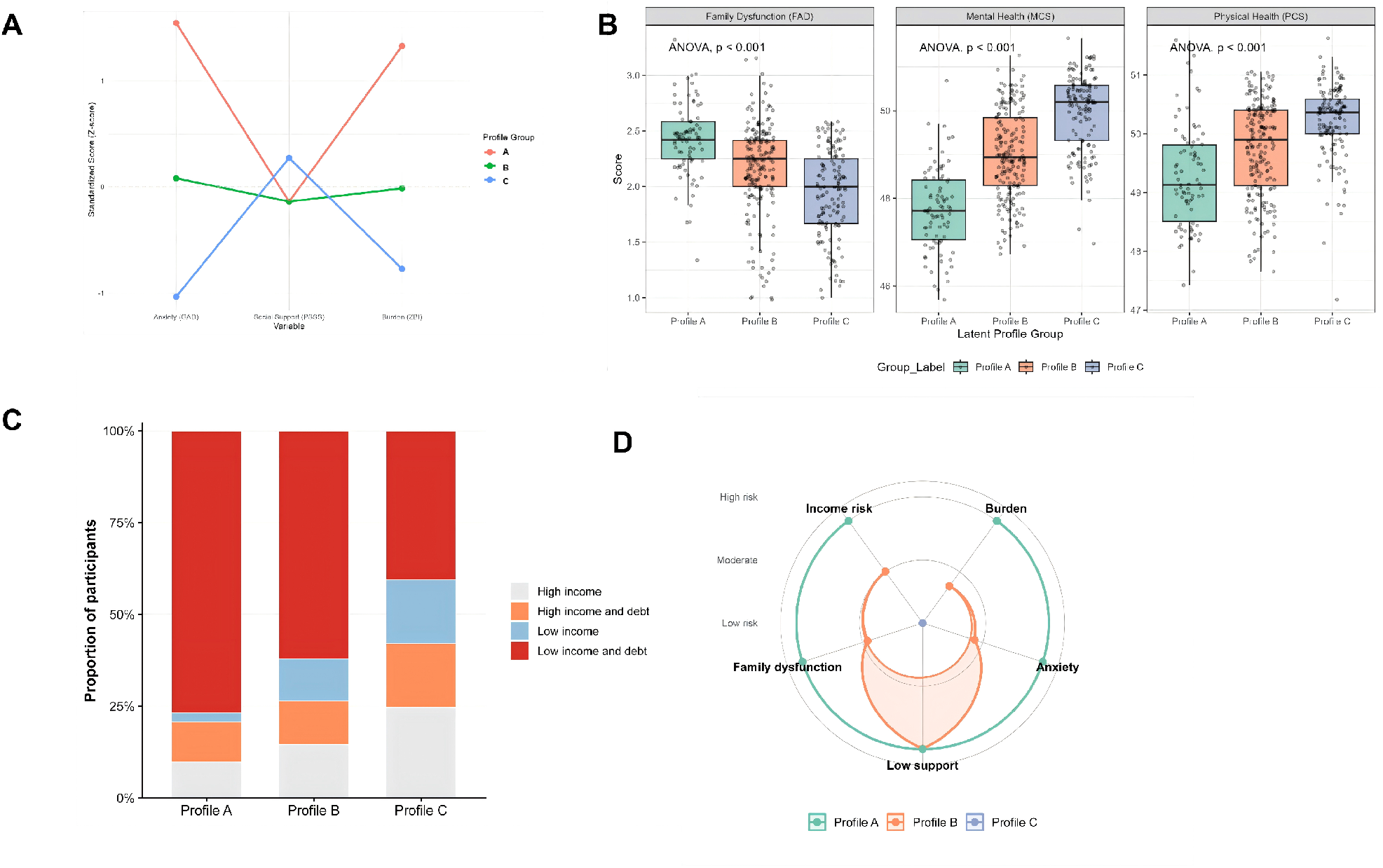
Caregiver profiling and clinical risk stratification by latent profile analysis. a) LPA identifying three caregiver profiles from standardized anxiety (GAD-7), perceived social support (PSSS), and caregiver burden (ZBI-22) scores. Profile A represents severe burden-high vulnerability, Profile B represents moderate burden, and Profile C represents mild burden-high resilience. Profile colors should be interpreted using the legend within each panel. b) Boxplots showing differences across profiles in family dysfunction (FAD-GF), mental quality of life (SF-MCS-12), and physical quality of life (SF-PCS-12). All outcomes differed across profiles (P < 0.001), and individual observations are overlaid. c) Stacked bar chart showing the distribution of combined income-debt categories within each profile. Profile A had the largest proportion of lower-income households with debt. d) Radar chart summarizing income risk, burden, anxiety, low social support, and family dysfunction across the three profiles. Concentric rings indicate increasing risk. Multinomial logistic regression predictors for profile membership are reported in Table 3.

The profiles also differed in outcomes not used to derive profile membership (Figure 3B). FAD-GF, SF-MCS-12, and SF-PCS-12 scores differed across profiles (all P < 0.001), with Profile A showing the least favorable mean values.

**Table 3.** Multinomial logistic regression analysis predicting latent profile membership. Profile C (mild burden-high resilience) was the reference category. Adjusted ORs and 95% CIs are reported. The model included child gender, disease laterality, family debt, and monthly household income. B, log-odds estimate; SE, standard error; OR, odds ratio.

| Table 3: Multinomial Logistic Regression Analysis for Latent Profile Groups |  |  |  |  |  |
| --- | --- | --- | --- | --- | --- |
| Variables | Estimate (B) | SE | Wald $\chi^2$ | P value | OR (95% CI) |
| Panel: Panel A: Moderate Caregiver Burden (Profile B) vs. Mild Caregiver Burden (Profile C) |  |  |  |  |  |
| Child Gender (Ref: Boy) |  |  |  |  |  |
| Girl | 0.42 | 0.21 | 4.08 | 0.043 | 1.52 (1.01, 2.29) |
| Onset Type (Ref: Unilateral) |  |  |  |  |  |
| Bilateral | 0.33 | 0.15 | 4.93 | 0.026 | 1.39 (1.04, 1.86) |
| Family Debt (Ref: Yes) |  |  |  |  |  |
| No | -0.46 | 0.20 | 5.36 | 0.021 | 0.63 (0.43, 0.93) |
| Monthly Income (Ref: High Income) |  |  |  |  |  |
| Low Income / Moderate Income | 1.85 | 0.55 | 11.32 | < 0.001 | 6.36 (2.16, 18.68) |
| Panel B: Severe Caregiver Burden (Profile A) vs. Mild Caregiver Burden (Profile C) |  |  |  |  |  |
| Child Gender (Ref: Boy) |  |  |  |  |  |
| Girl | 0.85 | 0.30 | 8.02 | 0.005 | 2.34 (1.30, 4.21) |
| Onset Type (Ref: Unilateral) |  |  |  |  |  |
| Bilateral | 0.67 | 0.22 | 9.25 | 0.002 | 1.95 (1.26, 3.02) |
| Family Debt (Ref: Yes) |  |  |  |  |  |
| No | -1.20 | 0.35 | 11.75 | < 0.001 | 0.30 (0.15, 0.60) |
| Monthly Income (Ref: High Income) |  |  |  |  |  |
| Low Income / Moderate Income | 3.45 | 0.80 | 18.60 | < 0.001 | 31.50 (6.56, 151.24) |

Multinomial logistic regression identified factors associated with profile membership (Table 3; Figure 3C,D). Compared with Profile C, low-to-moderate household income was associated with substantially higher odds of Profile A membership (OR = 31.50, 95% CI [6.56, 151.24], P < 0.001). The presence of family debt was associated with 3.33-fold higher odds of Profile A membership (OR = 3.33, 95% CI [1.67, 6.67], P < 0.001). Bilateral disease (OR = 1.95, 95% CI [1.26, 3.02], P = 0.002) and female child gender (OR = 2.34, 95% CI [1.30, 4.21], P = 0.005) were also associated with Profile A membership.

## Discussion

This cross-sectional study yielded three main findings. First, anxiety and socioeconomic hardship showed stronger associations with caregiver burden than the clinical indicators examined. Second, family functioning statistically accounted for approximately one-third of the association between perceived social support and caregiver burden, while household income modified the association between family dysfunction and burden. Third, nearly one in five caregivers belonged to a severe-burden, high-vulnerability profile, and lower household income and family debt were strongly associated with membership in this group.

Family functioning partly accounted for the inverse association between perceived social support and caregiver burden. This pattern is consistent with the stress-buffering hypothesis [24] and suggests that external support may be linked to caregiver well-being partly through family communication, role organization, and emotional responsiveness. Because the data are cross-sectional, the direction and temporal ordering of these associations cannot be established. Nevertheless, the findings support prospective evaluation of interventions that combine practical support with family-centered components.

The interaction between income and family dysfunction indicated greater socioeconomic vulnerability among lower-income households. This pattern is consistent with research on financial hardship during pediatric cancer treatment [25]. Direct medical expenses, nonmedical costs, employment disruption, and income loss may compound caregiving demands. The stronger economic associations during the initial-diagnosis phase further suggest that early financial screening warrants prospective evaluation within RB care pathways.

The profile analysis also showed that mean burden can conceal a highly distressed subgroup. The cohort mean (26.50 +/−15.02) was lower than values reported for cerebral palsy [6], mixed chronic pediatric conditions [26], and pediatric leukemia [7]. However, Profile A included 19.85% of caregivers and had a mean ZBI-22 score of 46.41, similar to the value reported among uninsured caregivers of children receiving active cancer treatment [9]. Burden among caregivers of children with celiac disease also increased markedly during the COVID-19 pandemic [27], illustrating how external stressors can intensify caregiver burden across pediatric conditions.

RB caregiving may involve a distinctive combination of concerns, including possible vision loss, enucleation, repeated examinations under anesthesia, and prognostic uncertainty. The present findings suggest that these clinical demands intersect with anxiety, family functioning, and financial conditions. However, the cross-sectional design does not establish a stress-sensitization mechanism or validate profile-specific interventions. The profiles should therefore be regarded as a screening framework for further evaluation rather than as a definitive clinical triage tool.

Several limitations should be considered. The cross-sectional design precludes causal inference and does not establish temporal ordering among burden, anxiety, family functioning, and economic status.

Participants were recruited from one tertiary ophthalmic oncology center, which may limit generalizability to other healthcare systems and resource settings. Self-reported measures may be affected by recall and social-desirability bias. Stepwise model selection may also produce unstable estimates, and the profiles require replication in independent cohorts. Longitudinal, multicenter studies should incorporate objective economic indicators, multi-informant family assessments, and prospective evaluation of profile-informed support.

In conclusion, caregiver burden in pediatric RB was associated with a combination of psychological, family, and socioeconomic factors. Anxiety had the strongest independent association with burden, family functioning partly accounted for the association between social support and burden, and household income modified the association between family dysfunction and burden. The severe-burden profile identified a subgroup that may warrant closer psychosocial and financial assessment. Longitudinal validation is needed before these profiles are used to allocate clinical interventions.

## Contributors

Peipei Zhang conceived and designed the study, developed the methodology, recruited participants, collected and curated the data, performed the statistical analyses, interpreted the findings, prepared the figures and tables, and drafted the manuscript. Xiaohua Ge supervised the study, contributed to the study design and interpretation of the findings, provided critical revision of the manuscript for important intellectual content, and oversaw the submission process. Both authors reviewed and approved the final manuscript and agree to be accountable for all aspects of the work. Xiaohua Ge is the guarantor and accepts full responsibility for the overall content, had access to the data, and controlled the decision to publish.

## Data availability statement

Deidentified participant-level data and the statistical code supporting the findings of this study are available from the corresponding author upon reasonable request, subject to approval by the institutional ethics committee and applicable data-protection requirements. Requests should include a methodologically sound research proposal and a data-use agreement.

## Ethics approval

This study involved human participants and was approved by the Ethics Committee of [Xinhua Hospital Affiliated to Shanghai Jiaotong University School of Medicine] (approval number: [XHEC-D-2026-048]).All participants provided written informed consent before participation.

