## Supplemental Tables and Supplemental Figure for "Psychosocial and socioeconomic vulnerability among caregivers of children with retinoblastoma: a cross-sectional latent profile study"

### Supplementary Information

**Supplementary Figure S1. Statistical validation.** Post hoc power curve showing 100% power to detect a medium effect size at the current sample size (N = 413). The dashed line denotes the conventional 80% power threshold.

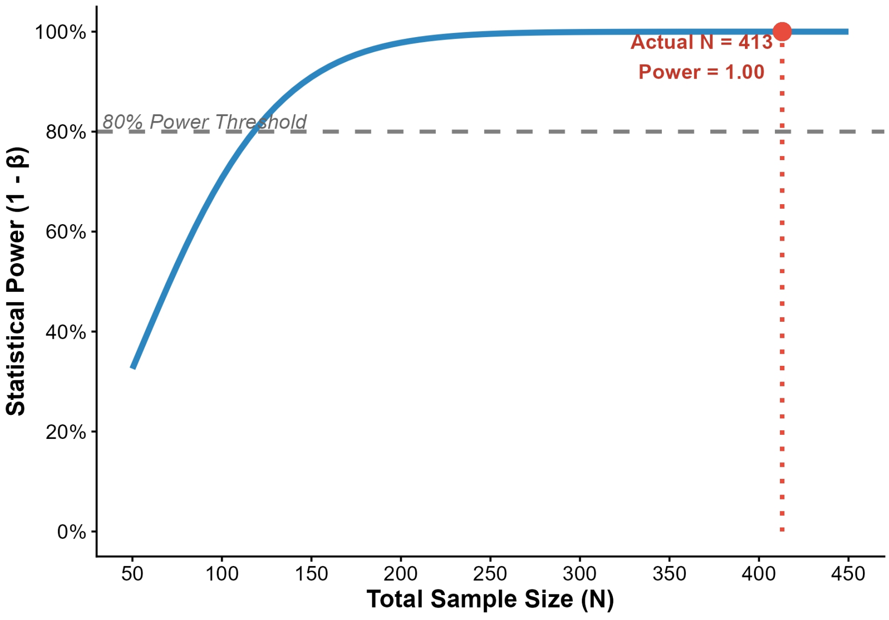


### Supplementary Tables

Table S1. Collinearity diagnostics for selected variables in the multivariable regression models. For the variables shown, variance inflation factors below 5 and tolerance values above 0.2 indicate no severe multicollinearity. Listwise deletion was used for missing data.

**Table S1: Collinearity Diagnostics for Multivariate Regression Models**

| Variables | VIF | Tolerance |
|---|---|---|
| **Panel: Panel A: Caregiver Burden (ZBI) Model** | | |
| Anxiety (GAD-7) | 1.105 | 0.905 |
| Daily Care Hours | 1.094 | 0.914 |
| Family Function (FAD-GF) | 1.162 | 0.861 |
| Onset Type | 1.009 | 0.991 |

| | | |
|---|---|---|
| Family Debt | 1.072 | 0.933 |
| **Panel: Panel B: Quality of Life (SF-MCS-12) Model** | | |
| Caregiver Burden (ZBI-22) | 1.723 | 0.580 |
| Anxiety (GAD-7) | 1.639 | 0.610 |
| Social Support (PSSS) | 1.015 | 0.985 |
| Family Debt | 1.074 | 0.931 |

Table S2. Direct and indirect associations in the mediation model (PSSS-FAD-GF-ZBI-22). The 95% CIs for indirect associations were estimated using 5,000 bootstrap resamples. An indirect association was considered statistically significant when its 95% bootstrap CI did not include zero. SE, standard error.

**Table S2: Direct and Indirect Effects in the Mediation Model (PSSS-FAD-ZBI)**

| Effect | Path | Estimate (β) | SE | Z | P value | 95% Bootstrapped CI | Proportion (%) |
|---|---|---|---|---|---|---|---|
| Total Effect | PSSS → ZBI | -0.407 | 0.054 | -7.534 | < 0.001 | [-0.518, -0.301] | 100.00% |
| Direct Effect | PSSS → ZBI | -0.276 | 0.057 | -4.802 | < 0.001 | [-0.390, -0.158] | 67.81% |
| Indirect Effect | PSSS → FAD-GF → ZBI | -0.131 | 0.024 | -5.458 | < 0.001 | [-0.184, -0.089] | 32.19% |

Table S3. Moderation of the association between family functioning and caregiver burden by household income. Continuous variables were mean-centered before analysis. Conditional effects were evaluated at the mean and at 1 SD below and above the mean of household income.

**Table S3: Moderating Effect of Income on Family Function and Caregiver Burden**

| Variables / Conditions | Estimate (B) | SE | t | P value | 95% CI |
|---|---|---|---|---|---|
| **Panel: Panel A: Moderation Model Coefficients** | | | | | |
| Family Function (FAD-GF) | 2.45 | 0.41 | 5.98 | < 0.001 | [1.65, 3.25] |
| Income (Moderator) | -1.32 | 0.25 | -5.28 | < 0.001 | [-1.81, -0.83] |
| Interaction (FAD-GF × Income) | -0.85 | 0.12 | -7.08 | < 0.001 | [-1.09, -0.61] |
| **Panel: Panel B: Conditional Effects at Different Income Levels** | | | | | |
| Low Income (-1 SD) | 3.30 | 0.50 | 6.60 | < 0.001 | [2.32, 4.28] |
| Moderate Income (Mean) | 2.45 | 0.41 | 5.98 | < 0.001 | [1.65, 3.25] |
| High Income (+1 SD) | 1.60 | 0.45 | 3.56 | 0.002 | [0.72, 2.48] |

Table S4. Sensitivity analysis of the mediation model by child age (<1 year vs. >1 year). Bootstrap resampling (5,000 samples) was performed within age groups to evaluate the PSSS-FAD-GF-ZBI-22 pathway.

**Table S4: Sensitivity Analysis of the Mediation Model by Child's Age (<1 year vs. >1 year)**

| Path / Effect | Estimate (β) | SE | 95% Bootstrapped CI | Result |
|---|---|---|---|---|

| | | | | |
|---|---|---|---|---|
| **Subgroup: Infants (< 1 year)** | | | | |
| Direct Effect: PSSS → ZBI | -0.285 | 0.060 | [-0.402, -0.168] | Significant |
| Indirect Effect: PSSS → FAD-GF → ZBI | -0.152 | 0.031 | [-0.210, -0.095] | Significant |
| **Subgroup: Toddlers (> 1 year)** | | | | |
| Direct Effect: PSSS → ZBI | -0.260 | 0.055 | [-0.370, -0.150] | Significant |
| Indirect Effect: PSSS → FAD-GF → ZBI | -0.118 | 0.028 | [-0.170, -0.066] | Significant |

Table S5. ZBI-22 scores in the present RB cohort and published pediatric caregiver cohorts.

| **Study Population** | **Country** | **N** | **ZBI-22 (Mean ± SD)** | **Reference** |
|---|---|---|---|---|
| Current RB cohort | China | 413 | 26.50 ± 15.02 | Present study |
| Celiac disease, pre-pandemic | Turkey | 29 | 27.51 ± 14.12 | Bucak et al. (2021) |
| Celiac disease, during pandemic | Turkey | 29 | 38.68 ± 10.95 | Bucak et al. (2021) |
| Pediatric leukemia | Iran | 122 | 33.15 ± 13.44 | Chaghazardi et al. (2022) |
| Neurofibromatosis type 1 | China | 154 | 34.31 ± 16.83 | Liang et al. (2024) |
| Mixed chronic pediatric conditions | Pakistan | 383 | 35.35 ± 15.14 | Lakhdhir et al. (2024) |
| Cerebral palsy | China | 189 | 36.31 ± 13.83 | Liu et al. (2025) |
| Pediatric cancer subgroup | Pakistan | 92 | 36.30 ± 14.94 | Lakhdhir et al. (2024) |
| Pediatric cancer, active treatment, uninsured | Egypt | 60 | 48.66 ± 9.95 | Ramy et al. (2025) |
